# How does CBT work? Causal discovery modelling locates the mechanisms of change in cognitive behavioural treatment for eating disorders

**DOI:** 10.64898/2026.07.21.26358524

**Authors:** Sarah Barakat, Mathew Varidel, Patrick Eades, Victor An, Ian Hickie, Frank Iorfino, Sarah Maguire

## Abstract

**Background:** Cognitive behavioural therapy (CBT) is the most frequently used and recommended therapy program for mental health, conditions, including for eating disorders. Despite decades of use, little is known about how change is produced in CBT across mental illnesses, and there has been no empirical validation of the causal structure of change for CBT treatment of eating disorders. Progress in optimisation of CBT and the development of alternatives particularly for non-responders, has been constrained by a lack of understanding of the mechanisms through which the therapy works. This study aimed to investigate, for the first time, mechanism by mechanism the causal direction of change across a therapeutic CBT package.

**Methods:** Directed acyclical graphs (DAGs) were estimated using causal discovery machine learning methods applied to data from a randomised controlled trial of a digital CBT program in 114 participants with bulimia nervosa. Eating disorder symptoms and psychological distress were assessed at each treatment session across the 10-session intervention in both conditions. Treatment effects were then estimated using the inferred causal structure.

**Results:** The inferred causal structure estimated direct effects of treatment on eating restraint (small to medium), alongside indirect effects upon preoccupation with eating, fear of weight gain, desire for weight loss, binge days, loss of control, over-exercise and fasting (small to medium). Eating restraint emerged as to the first and only node that treatment directly affected, suggesting that it is the primary mechanism through which treatment produces subsequent change in other behavioural and cognitive symptoms.

**Conclusion:** These findings provide the first empirical evidence, beyond cross-sectional or correlational analyses, for the long theorised mechanisms underlying CBT for eating disorders. Contrary to the conceptual model which posits CBT acts directly on dietary restraint and shape and weight concerns, our findings suggest treatment acts primarily through reductions in eating restraint, with downstream effects on shape- and weight-related concerns and other symptoms. To build our understanding of the processes by which effective treatments for eating disorders operate, progress personalised medicine, and optimise therapeutic outcomes, the methodology developed here can be applied broadly to examination of causal change structures in both direction and magnitude across different interventions.

## 1. Introduction

Despite being over fifty years old, and the most researched and prescribed psychotherapy in mental health, little is known about the mechanism of action of cognitive behavioural therapy (CBT). Meta-analyses across mental health consistently demonstrate large symptom reduction in both the short term and long term for CBT, and full remission in up to 20-45% of cases (1). In eating disorders the case is no different; CBT has the largest evidence-base, produces effect sizes on eating disorder symptomatology between 0.62x-2.75x (cohens *d*) and is prescribed as a first-line treatment for eating disorders, especially those marked by binge eating (2,3). Yet high rates of variability in treatment response persist. A significant proportion of people do not experience meaningful improvement after completing an evidence-based dose of CBT, and among those who do recover, up to 50% will relapse (4,5). Concerningly, research to date has failed to identify why treatment is ineffective for some, in part due to a limited understanding of how CBT brings about change in those who do respond. To optimise treatments for non-responders, it is essential to understand the *causal mechanisms*; that is the change processes through which treatment operates. This is a critical step in determining *why* treatments are effective for some people but not for others, and critically, *for whom* they are effective and for whom they are not (6,7).

The theoretical model underpinning CBT in general was formulated in the 1960s by Aaron T Beck (8) and quickly proliferated across mental health (9). Four decades ago the first specialised version of CBT for eating disorders was developed and trialled in bulimia nervosa (10), then expanded to CBT treatment for anorexia nervosa (11,12), binge eating disorder (11,13), and subsequently has been applied to almost every eating disorder diagnostic category (ARFID, (14); Body Dysmorphia, (15). The model purports to rely on two theoretical causal mechanisms: reducing dietary restraint and modifying shape and weight concerns (11). The first mechanism targeted in the treatment is dietary restraint; the hypothesis being that establishing a regular eating pattern will reduce dietary restraint, which in turn decreases biological hunger and urges to binge eat. The second proposed and targeted mechanism is reliance on shape and weight as the primary source of self-worth. The CBT theory posits that this is the central underlying driver of illness, or the core psychopathology of an eating disorder, and that fundamentally it drives engagement in disordered eating behaviours such as caloric restriction and strict adherence to dietary rules, which then results in binge eating and purging behaviours (11).

However, this conceptual model of CBT has not been empirically demonstrated. While association studies have found changes in dietary restraint and shape and weight concerns alongside changes in core eating disorder symptoms (binge eating, purge behaviours), these studies have largely relied on cross sectional or pre-post datasets that do not speak to causality (16). Moreso, the methods applied to these datasets typically assume single mediator or moderator associations, ignoring the complex interplay of behavioural, cognitive and emotional symptoms involved in illness and recovery (17).

Bayesian networks, falling under the umbrella of causal discovery analysis (CDA), are an alternative approach to learn the potential causal pathways between symptoms and outcomes (18–20). Directed acyclical graphs (DAGs) is a Bayesian approach which applies machine learning to a dataset to learn the probable dependency structure between variables, trying different causal structures and estimating their likelihood, until it determines the most probable causal structure (6). DAGs depict the data, representing both effects and their probable causal direction, represented as arrows between symptoms. CDA has recently been applied to the learning of individual-level causal pathways for those with bulimia nervosa (21), PTSD (22), alcohol use (23,24), schizophrenia (25) and general mental ill health (26). However, it has not been applied to the understanding of illness features, and treatment interactions, to yield insights into how therapy works.

In the current study, we applied Bayesian network analysis in the form of DAGs to a longitudinal, multiple time-point data set monitoring symptom change throughout CBT treatment for eating disorders marked by binge eating. While traditional statistical analysis of the treatment outcomes evaluated under our randomised controlled trials (RCT) determined an abstinence rate at follow-up of approximately 30% following CBT treatment and an effect size of up to 2.13 (27), these statistical approaches are unable to yield insights into how change within therapy occurs, nor what drives outcomes; knowledge which is vital for optimising delivery of treatment to improve abstinence, remission rates and outcomes in general.

### 1.1. Aim

We aimed to investigate whether the theoretically defined mechanisms of CBT for eating disorders can be empirically validated via exploration of the probable causal structures underpinning the session-by-session treatment effects of CBT in a sample of people with bulimia nervosa (see Barakat et al., 2021 (27) for full description of the RCT study). To contextualise these analyses, we first applied regression models to characterise change in global and item-level scores on measures of psychological distress and eating disorder symptoms administered at the beginning of every session. Then, using causal discovery analysis, we aimed to investigate though what purported mechanisms of CBT exerts its therapeutic effects.

## 2. Methods

### 2.1. Participants and Procedure

This study represents an exploratory, secondary analysis of a randomised controlled trial of an online 10-session CBT intervention for bulimia nervosa (*Binge Eating eTherapy* or *BEeT*). The study methodology is described in detail in the study protocol (28) and the primary outcomes of the RCT have been published (26). Participants (*N*=114) were recruited between May 2020 and October 2021 from the general Australian community, most commonly via referral from health professionals or advertisements on health websites and social media. Eligible participants were 16 years or older, met criteria for DSM-5 diagnosis of bulimia nervosa or subclinical bulimia nervosa (other specified feeding or eating disorder [OSFED] with bulimia behaviours) (30), had a body mass index (BMI) of 20 or above and were medically stable (see Barakat et al., 2021 for more details regarding eligibility criteria) (28).

After providing written informed consent, eligible participants were randomly assigned to one of three trial arms: 1) BEeT as a self-help programme (with administrative researcher contact), 2) BEeT with structured clinician-support (weekly 30-min telemedicine sessions), or 3) no-treatment (waitlist control; WLC).

All assessments were administered as online self-report measures. Assessments were completed at baseline, post-treatment (12 weeks after baseline) and 3-months after the post-treatment assessment, with a weekly symptom monitoring questionnaire completed at the start of each online session. The trial was pre-registered with the Australia New Zealand Clinical Trials Registry (ANZCTR Registration Number: ACTRN12619000123145p) and received ethical approval by the Royal Prince Alfred Hospital Human Research Ethics Review Board (HREC #X18–0486).

### 2.2. Measures

The Eating Disorder Examination-Questionnaire (EDE-Q) (31) was used to examine eating disorder symptomology, including frequencies of disordered eating behaviours (binge episodes, self-induced vomiting, laxative and diuretic use, driven exercise) over the preceding 28 day period, and eating disorder psychopathology according to four subscales (weight concern, shape concern, dietary restraint) and a global score (range: 0-6; higher scores represent greater eating disorder symptom severity; Cronbach’s a = 0.82). The EDE-Q was administered at baseline, post-treatment and follow-up.

A shorter 12-item version of the EDE-Q, the Eating Disorder Examination-Questionnaire Short (EDE-QS) (32), was administered at the beginning of each session during the intervention period. Using a 4-point Likert response scale, the EDE-QS assesses the severity of eating disorder behaviours and psychopathology theorised to be mechanistic for CBT (e.g. eating restraint, shape & weight concerns) in the previous seven day period. The EDE-QS total score is the sum of all responses (range: 0-36; higher scores represent greater eating disorder symptom severity; Cronbach’s a = 0.84).

The 10-item Kessler Psychological Distress Scale (K10) (33) was administered at baseline, post-treatment and follow-up to assess levels of psychological distress in the preceding four week period (range: 0-50; higher scores represent greater psychological distress). An adapted version of the K10 was administered per session, with the reference period adjusted to be the previous *week*, instead of the previous *four weeks*.

### 2.3. Statistical Analysis

Estimating total treatment effects were completed using the intention-to-treat principle. Missing data was dealt with using multiple imputation by chained equations (MICE) (34). We created five completed data sets via imputations each with 50 iterations. Convergence of the chains for the mean and standard deviation for each parameter under imputation was checked by-eye. For the total scores the predictor variables for imputations were baseline EDE-Q global score, K10 total score, age, illness duration, and gender. For the items, we used the factor-level scores relating to eating restraint, eating concern, shape concern, weight concern, as well as the K10 total score, age at presentation, illness duration, and gender. Data was not imputed for participants where there is incomplete data at baseline presentation (n=5).

#### 2.3.1. Regression Analysis

Linear regression was used to predict weekly outcomes. Outcomes were standardised by subtracting the population mean and standard deviation prior to performing the linear regression. We incorporated random slopes and intercepts nested per trial-arm. Pretreatment covariates of age, illness duration, gender, EDE-Q global, and K-10 total score were included in the regression. Posterior sampling was performed for each imputed data set using the Bayesian Regression Models using Stan package (34, 35). We ran the sampling procedure on each data set for 10000 iterations for two chains per imputed data set. Convergence statistics and resolution were checked for each imputed data set (max(R_hat) < 1.01, min(n_eff) > 400) (36, 37). Posterior samples across the imputed data sets were pooled and summarised. We report the estimated difference by the final session using the linear regression analysis, which is equivalent to a Cohen’s d estimand. Qualitative adjectives used to describe the absolute effect size point estimate (|*d*|) is as follows (39); negligible: |*d*| ≤ 0.01, very small: |*d*| ∈ (0.01, 0.20], small: |*d*| ∈ (0.20, 0.50], medium: |*d*| ∈ (0.50, 0.80], large: |*d*| ∈ (0.80, 1.20] and very large: |*d*| > 1.20. Credible evidence of improvement was defined as the 95% equal-tailed credible interval (CI) is inconsistent with zero.

#### 2.3.2. Causal Discovery Analysis

Causal discovery analysis was performed to learn the cumulative effects from pre-treatment to a given week. We assumed that the treatment variable and pre-treatment eating disorder items were root nodes. This provides a mechanism to account for baseline presentation. The dependency structure between the root nodes and items and between the items, at the analysed weeks was allowed to vary.

We performed two analyses. One to estimate the most probable DAG. In this case, we estimated this from a summarised imputed data set. Whereby, the multiple imputations were combined to take the average estimate per missing data point. We then ran tabu search 300 times starting from random locations (37, 38) and used the highest scoring structure using the Bayesian gaussian equivalent (BGe) score. The BGe score assumes that a target variable is regressed on parent nodes (i.e., nodes that directly point to it) assuming a linear regression with residuals that follow a normal distribution.

To estimate the uncertainty over the DAGs, we used a bespoke implementation of the Partition Markov chain Monte Carlo scheme (42, 43). We ran four chains per imputation for a total of 20 chains. Each chain was started from the most probable DAG given the imputed data set found by running tabu search 300 times starting from random locations. The chains were checked for convergence per imputed data set, where we ensured all R_hat < 1.10 and resolution per chain corresponded to (*n*_eff_ ≥ 100) for the log-posterior DAG scores. The Bayesian gaussian equivalent score was used to retain the ordinal information in the data. The posterior distribution for DAGs was estimated by pooling the posterior samples across imputed data sets. We summarise the posterior distribution using a ‘consensus graph’, which shows arrows where the probability is greater than 20%, and weights those arrows in accordance with the posterior edge probability.

#### 2.3.3. Treatment Effect Estimation

Causal discovery can be used to estimate treatment effects on eating disorder symptoms. The existence of a path from treatment to a symptom suggests that there is a treatment effect. The type of path also informs the causal process of the treatment. A direct effect on a symptom is identified when there is an arrow from treatment to that symptom. An indirect effect on a symptom is identified when there is a path (i.e., series of arrows) from the treatment node to that symptom. We use this information to estimate the direct, indirect, and total (i.e., direct + indirect) effects of the treatment. We report several indices including, the probability that there is an effect, the effect size given the most probable graph, and the effect size marginalised over the probability distribution of causal structures (i.e., after incorporating the uncertainty of the structure). We also highlight those where the posterior probability of their being a path between treatment and outcome is greater than 50%, which meets the threshold of the edge being more probable than not, is consistent with Bayesian prediction model selection (44) and has been applied in mental health analysis (45).

## 3. Results

### 3.1. Participants

A total of 114 participants provided informed consent and were randomised to the treatment group (self-help (*n*=38 [33%]; clinician-supported (*n*=37 [32%]) or WLC (*n*=39 [34%]). The mean age of participants was 31.1 years (SD=10.2; range 16.0-65.0), with a mean BMI of 25.7 (SD=6.0; range 19.1–48.4). See Table 1 for baseline characteristics.

**Table 1.**
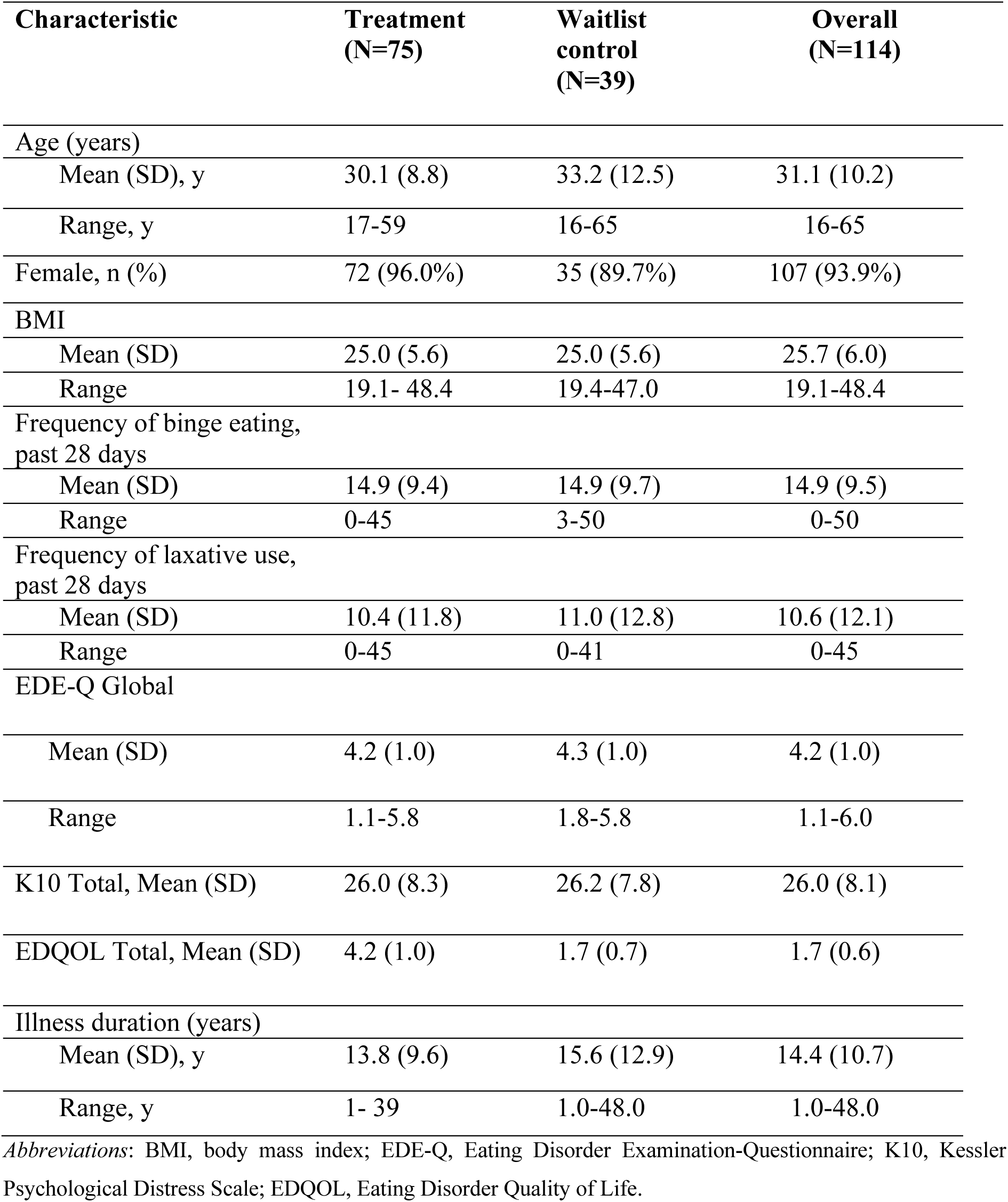
Baseline Characteristics of Participants.

| **Characteristic** | **Treatment**<br>**(N=75)** | **Waitlist**<br>**control**<br>**(N=39)** | **Overall**<br>**(N=114)** |
| --- | --- | --- | --- |
| **Age (years)** |  |  |  |
| Mean (SD), y | 30.1 (8.8) | 33.2 (12.5) | 31.1 (10.2) |
| Range, y | 17-59 | 16-65 | 16-65 |
| Female, n (%) | 72 (96.0%) | 35 (89.7%) | 107 (93.9%) |
| **BMI** |  |  |  |
| Mean (SD) | 25.0 (5.6) | 25.0 (5.6) | 25.7 (6.0) |
| Range | 19.1- 48.4 | 19.4-47.0 | 19.1-48.4 |
| **Frequency of binge eating,**<br>**past 28 days** |  |  |  |
| Mean (SD) | 14.9 (9.4) | 14.9 (9.7) | 14.9 (9.5) |
| Range | 0-45 | 3-50 | 0-50 |
| **Frequency of laxative use,**<br>**past 28 days** |  |  |  |
| Mean (SD) | 10.4 (11.8) | 11.0 (12.8) | 10.6 (12.1) |
| Range | 0-45 | 0-41 | 0-45 |
| **EDE-Q Global** |  |  |  |
| Mean (SD) | 4.2 (1.0) | 4.3 (1.0) | 4.2 (1.0) |
| Range | 1.1-5.8 | 1.8-5.8 | 1.1-6.0 |
| K10 Total, Mean (SD) | 26.0 (8.3) | 26.2 (7.8) | 26.0 (8.1) |
| EDQOL Total, Mean (SD) | 4.2 (1.0) | 1.7 (0.7) | 1.7 (0.6) |
| **Illness duration (years)** |  |  |  |
| Mean (SD), y | 13.8 (9.6) | 15.6 (12.9) | 14.4 (10.7) |
| Range, y | 1- 39 | 1.0-48.0 | 1.0-48.0 |

*Abbreviations:* BMI, body mass index; EDE-Q, Eating Disorder Examination-Questionnaire; K10, Kessler Psychological Distress Scale; EDQOL, Eating Disorder Quality of Life.

### 3.2. Main Effects

### 3.2.1. Total Scores

Figure 1 depicts the change in EDE-QS and K10 total scores per-session across the 10-session intervention period. The treatment arms resulted in improved EDE-QS total scores (*d* = -0.951, 95% CI [-1.365, -0.537]), compared to WLC. Whereas the improvement in K10 items remained consistent with zero (*d* = -0.386, CI [-0.809, 0.069]).

**Figure 1.**
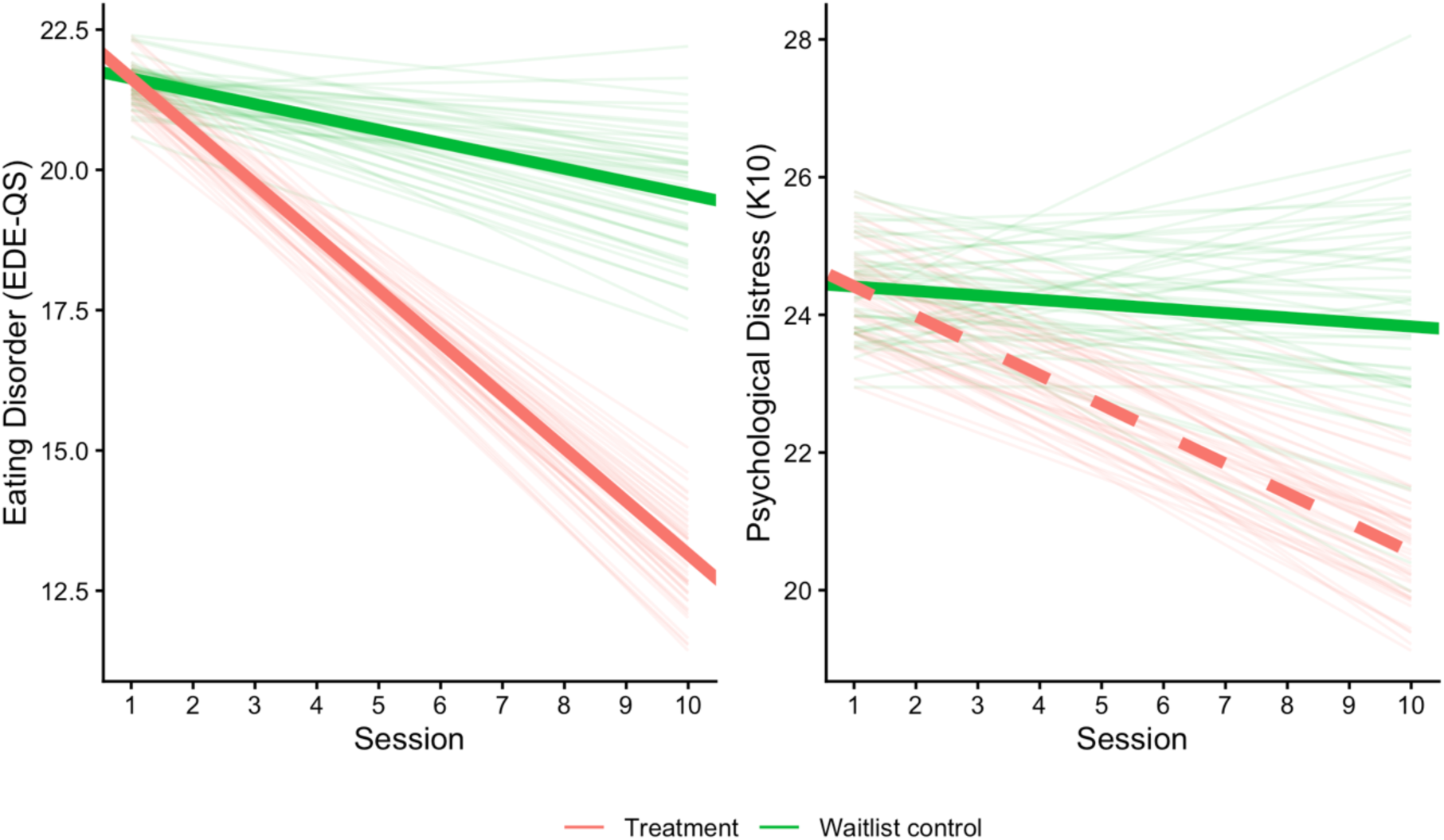
Linear Time Series Model of Per-Session Change in Total Scores of EDE-QS and K10 Note. Bold lines represent the group-specific linear regression model and lighter lines represent posterior samples of the linear regression model. The solid lines represent comparisons where the effect size of per-session change in the intervention arm is greater than WLC at 95% CI, whereas broken lines represent comparisons where the intervention arm does not outperform WLC at 95% CI. Abbreviations: WLC, EDE-QS, Eating Disorder Examination-Questionnaire Short; K10, Kessler Psychological Distress Scale.

#### 3.2.2. Item-Level Scores

Figure 2 compares the effect size by the final session for each item of the EDE-QS and K10. Several eating disorder items exhibit credible evidence of improvement within the treatment group compared to WLC. These include eating restraint (*d* = -0.626, CI [-0.985, -0.271]), fasting (*d* = -0.583, CI [-0.966, -0.199]), preoccupation with eating (*d* = -0.632, CI [-1.004 -0.270]), preoccupation with weight and shape (*d* = -0.457, CI [-0.860, -0.061]), fear of weight gain (*d* = -0.604, CI [-1.027, -0.158]), and perceived loss of control (*d* = -0.566, CI [-1.025, -0.094]), binge eating (*d* = -0.681, CI [-1.048, -0.283]). Two items relating to depressive symptoms in the psychological distress scale improved in the treatment group compared to WLC. These included tiredness (*d* = -0.390, CI [-0.765, -0.094]) and worthlessness (*d* = -0.335, CI [-0.638, -0.019]). Further numerical detail can be found in Supplementary Material.

**Figure 2.**
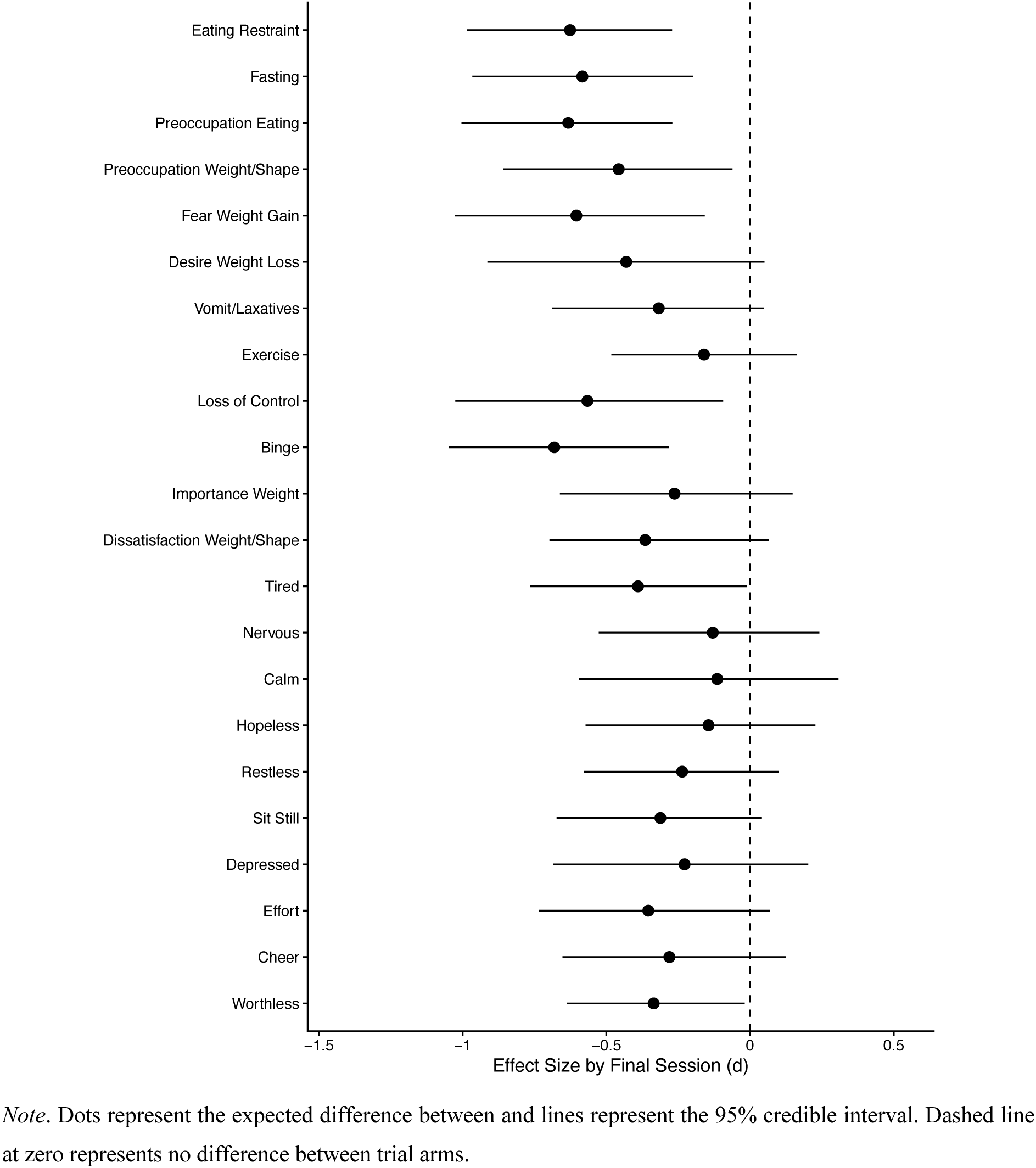
Comparison of Change in Item-Level Scores of EDE-QS and K10 Between Treatment and Control Conditions from Baseline to Post-Treatment Note. Dots represent the expected difference between and lines represent the 95% credible interval. Dashed line at zero represents no difference between trial arms.

**Figure 3.**
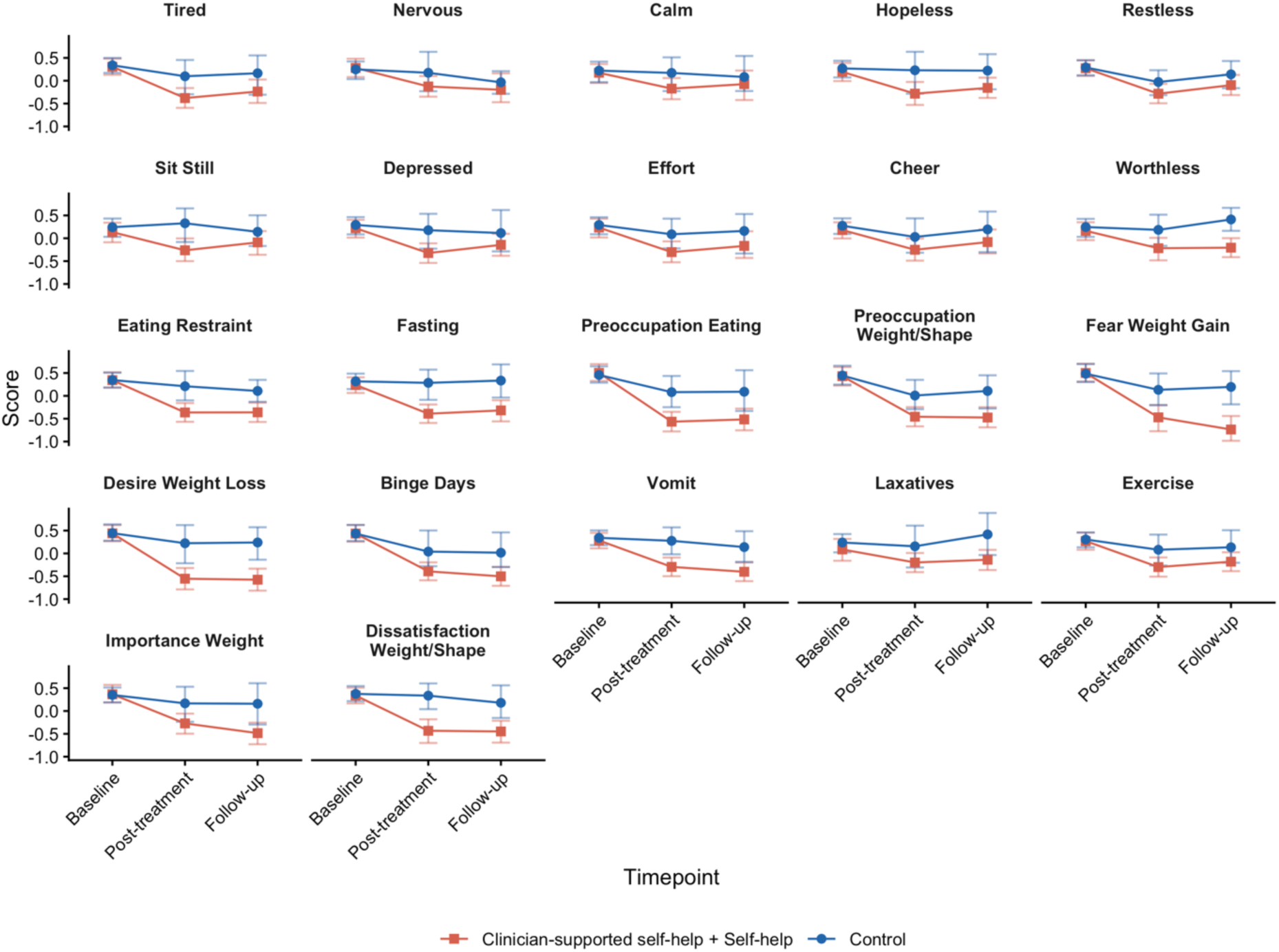
Item-Level Scores of K10 Across the 10 Sessions for each Condition at Baseline, Post-Treatment and Follow-up

We also checked the effects of CBT on individual symptoms at follow up. We found that most eating disorder items exhibited improvement at follow up, including; desire for weight loss (Δ = -0.772, CI [-1.285, -0.235]), dissatisfaction with weight/shape (Δ = -0.733, CI [-1.074, -0.436]), preoccupation with eating (Δ = -0.692, CI [-1.045, -0.361]), fear of weight gain (Δ [standardised difference] = -0.618, CI [-1.099, -0.185]), fasting (Δ = -0.596, CI [-0.906, -0.245]), eating restraint (Δ = -0.572, CI [-0.963, -0.223]), vomiting (Δ = -0.513, CI [-0.818, -0.200]), importance of weight (Δ = -0.453, CI [-0.814, -0.042]), and binge days (Δ = -0.434, CI [-0.886, -0.091]). Improvement on K10 items was only found for can’t sit still (Δ = -0.483, CI [-0.845, -0.096]) and feeling depressed (Δ = -0.428, CI [-0.782, -0.051]).

### 3.4. Causal Discovery Analysis

Results of the causal discovery analysis are shown graphically in Figures 4 and 5. Figure 4 shows the most probable causal structure, whereas Figure 5 shows the probable distribution of the causal structures.

**Figure 4.**
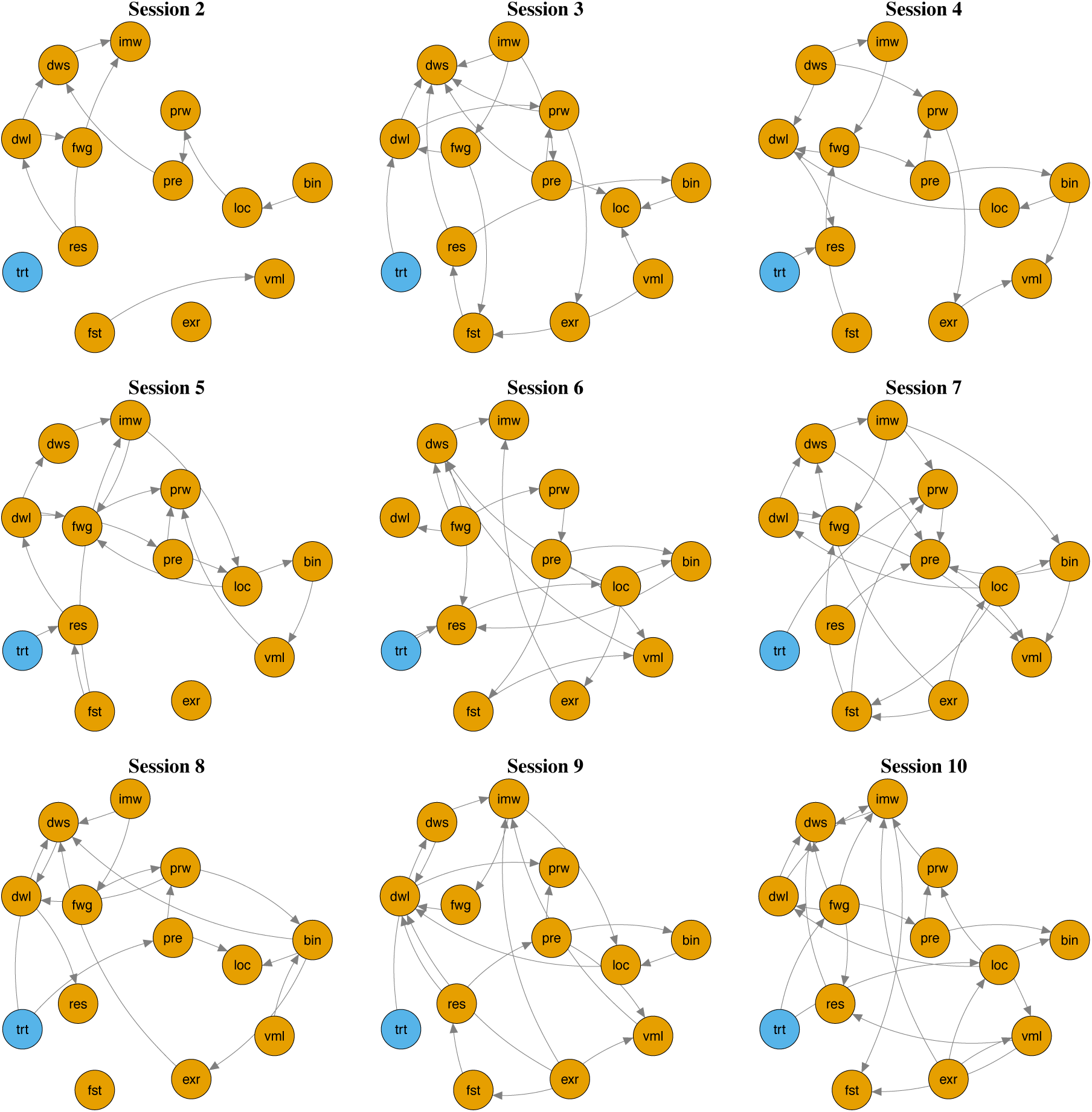
Representation of Most Probable Causal Paths between Treatment and Eating Disorder Symptoms per Session Across the Intervention Period Note. Nodes; trt: “Treatment”, res: “Eating Restraint”, fst: “Fasting”, pre: “Preoccupation Eating”, prw: “Preoccupation Weight/Shape”, fwg: “Fear Weight Gain”, dwl: “Desire Weight Loss”, vml: “Vomit/Laxatives”, exr: “Exercise”, loc: “Loss of Control”, bin: “Binge”, imw: “Importance Weight”, dws: “Dissatisfaction weight/shape”.

**Figure 5.**
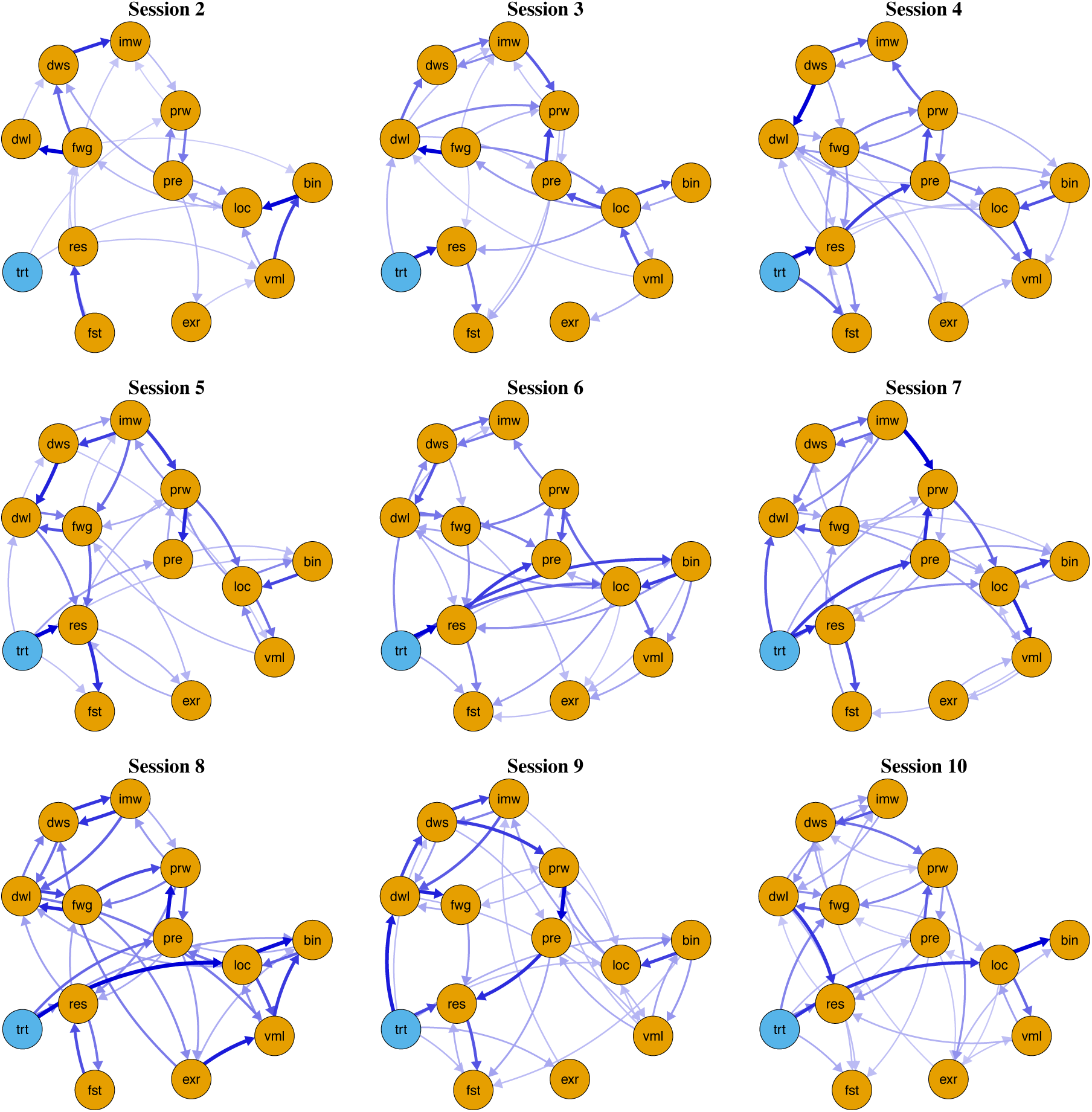
Consensus Graph Representation of Probable Distribution of Causal Structures per Session Across the Intervention Period Note. This figure summarises the posterior distribution of DAGs as marginalised edge probabilities where bolder edges represent greater probability. Nodes; trt: “Treatment”, res: “Eating Restraint”, fst: “Fasting”, pre: “Preoccupation Eating”, prw: “Preoccupation Weight/Shape”, fwg: “Fear Weight Gain”, dwl: “Desire Weight Loss”, vml: “Vomit/Laxatives”, exr: “Exercise”, loc: “Loss of Control”, bin: “Binge”, imw: “Importance Weight”, dws: “Dissatisfaction weight/shape”.

#### 3.4.1. Treatment Effects

The treatment most likely directly acts on eating restraint. This is due to an arrow appearing between treatment and eating restraint in the most probable causal structure by Sessions 3, 4, 5, 6, 7, 8, and 10. Correspondingly, uncertainty estimation suggests that treatment directly effects eating restraint with greater than 50% probability by Sessions 3 (p [probability of a direct causal influence], 52%), 4 (p, 82%), 5 (p, 77%), 6 (p, 94%), and 10 (p, 59%). Given the most probable structure, the direct effect on eating restraint by Session 10 was small (M, -0.38, IQR, -0.64, 0.00) to medium (mean, -0.58, SD, 0.28). These estimates alongside uncertainties can be found in Table 2.

**Table 2.** Total, Direct, and Indirect Effects of Treatment on EDE-QS for Session 10.

|  |  | Total Effects |  |  | Direct Effects |  |  | Indirect Effects |  |  |
| --- | --- | --- | --- | --- | --- | --- | --- | --- | --- | --- |
| Item | Name | MAP Estimate,<br>Mean (SD) | Overall Estimate,<br>Median (IQR) | P | MAP Estimate,<br>Mean (SD) | Overall Estimate,<br>Median (IQR) | P | MAP Estimate,<br>Mean (SD) | Overall Estimate,<br>Median (IQR) | P |
| 1 | Eating Restraint | -0.73<br>(0.28) | -0.58<br>(-0.74, -0.42) | 0.96<sup>†</sup> | 0.64<br>(0.15) | -0.26<br>(-0.54, 0.00) | 0.59<sup>†</sup> | -0.08<br>(0.23) | -0.30<br>(-0.53, -0.02) | 0.70<sup>†</sup> |
| 2 | Fasting | -0.62<br>(0.16) | -0.55<br>(-0.70, -0.40) | 0.94<sup>†</sup> | -0.54<br>(0.14) | 0.00<br>(-0.37, 0.00) | 0.40 | -0.07<br>(0.22) | -0.4<br>(-0.59, -0.13) | 0.74<sup>†</sup> |
| 3 | Preoccupation Eating | -0.52<br>(0.18) | -0.22<br>(-0.38, -0.06) | 0.96<sup>†</sup> | 0.00 | 0<br>(0.00, 0.00) | 0.10 | -0.52<br>(0.18) | -0.21<br>(-0.36, -0.04) | 0.92<sup>†</sup> |
| 4 | Preoccupation Weight/Shape | -0.41<br>(0.18) | -0.10<br>(-0.27, 0.03) | 0.94<sup>†</sup> | 0.00 | 0.00<br>(0.00, 0.00) | 0.06 | -0.41<br>(0.18) | -0.10<br>(-0.28, 0.03) | 0.91<sup>†</sup> |
| 5 | Fear Weight Gain | -0.49<br>(0.19) | -0.43<br>(-0.63, -0.21) | 0.92<sup>†</sup> | 0.00 | 0.00<br>(0.00, 0.00) | 0.16 | -0.49<br>(0.19) | -0.37<br>(-0.57, -0.13) | 0.87<sup>†</sup> |
| 6 | Desire Weight Loss | -0.47<br>(0.21) | -0.59<br>(-0.78, -0.36) | 0.90<sup>†</sup> | 0.00 | 0.00<br>(0.00, 0.00) | 0.08 | -0.47<br>(0.21) | -0.56<br>(-0.77, -0.30) | 0.88<sup>†</sup> |
| 7 | Vomit/Laxatives | -0.45<br>(0.17) | -0.16<br>(-0.31, 0.00) | 0.91<sup>†</sup> | 0.00 | 0.00<br>(0.00, 0.00) | 0.07 | -0.45<br>(0.17) | -0.15<br>(-0.31, 0.00) | 0.86<sup>†</sup> |
| 8 | Exercise | -0.21<br>(0.16) | -0.30<br>(-0.46, -0.08) | 0.86<sup>†</sup> | 0.00 | 0.00<br>(0.00, 0.00) | 0.08 | -0.21<br>(0.16) | -0.27<br>(-0.44, -0.03) | 0.81<sup>†</sup> |
| 9 | Loss of Control | -0.69<br>(0.18) | -0.43<br>(-0.57, -0.27) | 0.97<sup>†</sup> | -0.37<br>(0.13) | 0.00<br>(-0.33, 0.00) | 0.39 | -0.32<br>(0.23) | -0.28<br>(-0.48, -0.05) | 0.82<sup>†</sup> |
| 10 | Binge | -0.53<br>(0.18) | -0.32<br>(-0.47, -0.17) | 0.96<sup>†</sup> | 0.00 | 0.00<br>(0.00, 0.00) | 0.17 | -0.53<br>(0.18) | -0.28<br>(-0.44, -0.01) | 0.87<sup>†</sup> |
| 11 | Importance Weight | -0.33<br>(0.21) | -0.12<br>(-0.32, 0.00) | 0.77<sup>†</sup> | 0.00 | 0.00<br>(0.00, 0.00) | 0.12 | -0.33<br>(0.21) | -0.09<br>(-0.29, 0.00) | 0.69<sup>†</sup> |
| 12 | Dissatisfaction weight/shape | -0.45<br>(0.20) | -0.23<br>(-0.46, 0.00) | 0.84<sup>†</sup> | 0.00 | 0.00<br>(0.00, 0.00) | 0.12 | -0.45<br>(0.20) | -0.20<br>(-0.42, 0.00) | 0.79<sup>†</sup> |

*Note.* Dagger represents when the probability of a path is greater than 50% probability.

We found small to medium indirect effects of treatment on a range of variables by the final session, including eating restraint (M, -0.30, IQR, -0.53, -0.02), fasting (M, -0.40, IQR, -0.59, -0.13), preoccupation with eating (M, -0.21, -0.36, -0.04), fear of weight gain (M, -0.37, IQR, -0.57, -0.13), desire for weight loss (M, -0.56, IQR, -0.77, -0.30), over-exercising (M, -0.27, -0.44, -0.03), loss of control (M, -0.28, IQR, -0.48, -0.05), and binge days (M, -0.28, IQR, -0.44, -0.01). The paths by which CBT indirectly influences these symptoms are less clear. For example, examining the edge probabilities for session ten, the evidence for eating restraint acting on other nodes is limited. Specifically, eating restraint potentially influences desire for weight loss (p [probability of path], 0.37), use of vomit and laxatives (p, 0.27), fasting periods (p, 0.26), preoccupation with eating or weight/shape (p, 0.25), or over-exercising (p, 0.22), but with probability of these paths being less than 50%. In short, while the existence of an indirect path from treatment to these variables is more probable than not, and most probably occurs via eating restraint, there is uncertainty.

Several other interactions between eating disorder symptoms were identified across multiple sessions. Perceived loss of control tended to interact with use of vomit/laxatives by each session. Specifically, probability of an edge in either direction between use of vomit/laxatives and perceived loss of control was 97% by Session 3, 65% by Session 4, 95% by Session 5, 53% by Session 6, 58% by Session 7, 59% by Session 8, 43% by Session 9, and 99% by Session 10. Similarly, loss of control and binge eating tended to interact, with at least 90% probability of a direct interaction by each session. However, directionality between these symptoms tended to flip.

Also, cognitive factors such as the importance of weight and dissatisfaction with weight and/or shape showed an interaction across multiple sessions. The probability of a direct interaction between these variables was at least 82% by each session. Similarly, fear of weight gain and desire for weight loss tended to interact, with probability of a least 75% by each session. However, the directionality cannot be inferred from the data alone.

## 4. Discussion

In the current study, we applied causal discovery analysis to week-by-week symptom change data for a cohort of people with bulimia nervosa receiving CBT treatment for 10 weeks to empirically examine the theoretically proposed mechanisms of change in CBT. To our knowledge this is the first empirical examination in the literature of the purported mechanisms of change in CBT for these eating disorders, even though the treatment has been in use for almost 50 years. The probable causal models demonstrate that the CBT treatment most likely acts directly on eating restraint and indirectly upon preoccupation with eating, fear of weight gain, desire for weight loss, binge days, loss of control, over-exercise and fasting. The results of the regression analyses demonstrate that participants in the treatment arm outperform WLC in per-session change across majority of eating disorder symptoms. For depressive symptoms, there appeared to be a unique effect of treatment on symptoms including tiredness and worthlessness compared to WLC.

The results suggest that a wide range of paths both from treatment and between items are possible. However, certain trends exist. First, the networks become denser over sessions. Similar trends in the structural analysis have been found in previous research which has also applied a network approach to clinical trial data (46). This is likely due to investigating cumulative changes across the sessions. That is, the effects likely accumulate over multiple sessions, and thus evidence is only found for certain interactions at later sessions.

Second, our findings broadly support the theoretical underpinnings of the CBT model, demonstrating that treatment exerts its therapeutic effects through the reduction of dietary restraint and modification of shape and weight concerns (9, 42). However, our findings offer a unique insight into the temporal sequencing and association between these mechanisms, addressing key methodological shortcomings of previous research (48). Our results found eating restraint to be the first and only node that treatment *directly* affected, suggesting that it is the primary mechanism through which treatment produces change. The modelling explicates further our understanding of the relationship between key behaviours and features of an eating disorders, suggesting that eating restraint itself may be linked to and cause a number of the other distressing features of an eating disorder, which emerged as indirect effects, including desire for weight loss, self-induced vomiting and purging, fasting, overexercising and negative preoccupations with eating, weight and shape. Our findings suggest that the second proposed mechanism of CBT (modification of shape and weight concerns) operates first through changes in eating restraint. It may be that this sequencing simply reflects the structure and ordering of skills in CBT, whereby eating restraint is the first targeted mechanism through the introduction of self-monitoring and regular eating, followed by cognitive techniques designed to address weight and shape concerns (11). However, given that a direct effect of treatment upon weight and shape concerns was not identified in the subsequent weeks, it may be more likely that behavioural change in dietary restraint directly influenced related cognitive processes. For example, regular eating directly challenges fears regarding weight gain. This interpretation aligns with research more broadly on feared stimuli, in which the effects of repeated exposure to feared situations can facilitate the challenging of maladaptive beliefs through processes such as expectancy violation (49).

Third, several interactions between eating disorder symptoms themselves were identified in the modelling. These include the relationship between loss of control, vomiting/laxative use and binge eating; mapping closely onto the binge-compensation cycle embedded within the CBT model (45). The modelling showed that in Sessions 3 and 4, it appeared that loss of control was driving vomiting and laxative use behaviours, but Session 5 data suggested that might be that change in vomiting and laxative behaviours. It is not clear from the modelling to date whether a reduction in loss of control is needed in order to see a reduction in vomiting and laxative use behaviours or if the relationship may work the other way around. There are a number of possible explanations for this. Data limitations is one of them but it also may be that other mechanisms are involved in a dynamic and complex interaction requiring further explanation.

We also observed evidence of direct interactions between two key cognitive features of illness; the importance of weight or shape in self-image and dissatisfaction with weight or shape. These features are positioned as core drivers of eating disorder psychopathology in CBT theory, and yet our results found treatment to have a low or negligible effect on them in the network. Interestingly, treatment did appear to influence other cognitive features of illness, including preoccupation with eating, fear of weight gain and desire for weight loss. Differences in treatment responsiveness potentially suggests a multidimensional system of cognitive constructs, consisting of pervasive and deeply rooted negative beliefs about the self; distinct from ‘surface-level’ cognitions or attitudes related to one’s weight and eating, which appear to be more easily targeted. Indeed, this aligns with seminal cognitive models of psychopathology (46, 47) and is supported by empirical evidence showing that negative core beliefs in eating disorders are separate from, and predictive of, eating disorder cognitions and behaviours (50).

However, the CBT model of eating disorders clusters these constructs together under the banner of ‘over-evaluation of eating, shape and weight’ and accordingly most outcome studies of CBT similarly cluster these and do not distinguish between them (11). The clarification and detangling of mechanistic drivers within our most commonly used treatments, and data speaking to the differential impacts of care packages on some and not others is a vital step for treatment optimisation. Indications from this data, reflective of broader theory, suggest CBT treatment may benefit from the addition of therapeutic element targeting negative core beliefs about self and body.

Our results provide evidence consistent with a large body of research showing that early change within first four sessions of CBT is the most reliable predictor of treatment response (16). The strongest probabilities of direct effects on eating restraint occur within the corresponding timeframe between Session 4 (82%) and Session 6 (94%), indicating that eating restraint is likely to be the primary mechanism underlying early CBT response. Empirical support for the causal mechanisms likely to be responsible for early response offers a critical opportunity to meaningfully advance personalised care. By routinely measuring empirically identified early mechanisms of change such as dietary restraint at critical treatment points (e.g., sessions 4 and 6), clinicians can determine whether a patient is on the expected trajectory. When treatment response is not as expected, this can guide timely adjustments including intensifying or augmenting the treatment being delivered or pivoting towards an alternative treatment model to yield an optimised treatment response. Early treatment response is also a reliable predictor of CBT outcomes across several other mental health conditions (49, 50), suggesting the implications of this analytical approach may have broader applicability across psychiatry.

### Strengths & Limitations

To our knowledge, this study is the first to examine mechanism by mechanism the causal direction of change across a therapeutic CBT package. This study addresses key methodological limitations of prior work by applying a causal network model to longitudinal data with repeated symptom measurement at each treatment session, enabling estimation of the causal mechanisms of CBT for eating disorders for the first time. There are a number of studies that have attempted to examine the mechanisms of change across a therapeutic CBT package (51, 52), however for the most part the methodologies used select a priori a likely mediator of change and then estimate effect sizes (i.e. single mediator models). CDA, like all modelling, makes assumptions, however is able to take a more holistic approach; it has the capacity to understand multiple symptoms simultaneously and estimate the causal interactions between them. This method can be applied to datasets more broadly of CBT treatment effects, if regular measurement between delivery of therapeutic components is undertaken. Digital delivery of CBT in this study represents another key strength, providing embedded measurement capabilities and close monitoring of symptom change, alongside treatment delivered in a modular structure with high fidelity and no risk of therapist drift. This allows for precise examination of effects of specific treatment components on symptom change in such a way that is much more difficult with in-person formats.

Limitations exist in applying causal discovery analysis to mental health data. One that is applicable to our analysis, is that the model needs to assume that the underlying causal structure follows a DAG, which models relationships between symptoms as *acyclic,* the model struggles to deal with the fact that there is likely a bidirectional relationship between some symptoms. Trying to determine cyclic causal influence or bidirectional relationships between symptoms is difficult from observational data (55). There are other approaches that can be applied, such as directed cyclic graphs (56) or analyses that infer relationships between different timepoints (57, 58). However, all these methods make assumptions and therefore have limitations. While replication of these findings in a larger sample is desirable, large cohorts treated with discrete treatment components and repeated measures with high completion rates, is relatively rare in mental health. Further, the sample sizes required to estimate causal structures is not well understood (58). To address potential sample size limitations in this study, we conducted rigorous uncertainty estimates of the causal structure and the effects. Future work in this area is important to understand in particular the drivers of the poorest outcomes of CBT.

While the outcome measure used in this study (EDE-QS) enabled the examination of causal mechanisms, because it is a validated measure of the purported mechanisms in the CBT model, it must be noted that it does not include measurement of other key therapeutic processes described by both clinicians and lived experience as central to recovery, for example improved self-compassion, interoceptive awareness or reduced emotional avoidance (60). Future research including measurement of mechanisms not encapsulated in the CBT model is needed. Finally, while the CBT program delivered here is in weekly modules with measurement in between each, weekly sessions do not always focus on a single therapeutic skill with some sessions (particularly later in the 10 weeks) containing several skills, hence the potential impact of some components of therapy is not dealt with in this study (58, 59). To advance mechanism-informed treatments, future research should employ dismantling or factorial study designs that experimentally manipulate individual treatment components, thereby enabling direct evaluation of the causal mechanisms of each component of care.

### Clinical Implications

Our findings hold key implications for clinical care. They represent a critical step towards empirical understanding of which treatment components, delivered in what order, produce the best outcomes, rather than relying on clinical judgement alone. Network theory has gained prominence in recent years as a data-driven approach to personalised clinical care within psychiatry (63–66). Within this broader framework, CDA and DAGs extend network-based approaches by moving beyond estimation of symptom associations and towards the identification of potential causal relationships. Network models have been proposed as tools for identifying potential treatment targets and strengthening clinical case conceptualisations traditionally derived from clinician input alone. While the most commonly proposed approach is to use centrality indices (a measure of symptom association within a network) to identify treatment targets (40, 64–66), these indices are commonly derived from undirected networks that estimate associations rather than causal influence, and as a result, may not represent the most effective treatment targets (72). In contrast, the CDA approach used in this study employs DAGs identify potential causal pathways and estimate causal effects, providing greater insight into the directionality and magnitude of influence between symptoms, behaviours, and contextual factors

Causal effects provide the magnitude of influence, and can therefore be used to assign interventions, by comparing outcomes under different intervention scenarios to inform recommendations (19, 41). For example, individuals are suspected to have different causal processes that develop and sustain disorders, which will not be known *a priori* and thus learning those causal structures at the individual-level using an automated approach will likely be important. Such an approach could allow for automated treatment recommendations for an individual consistent with causal considerations. In order to optimise treatment efficacy, to progress personalised application of treatment elements and to understand therapeutic change in general that ability to examine whole treatment packages and both the direct and indirect effects of treatment mechanisms in interaction with each other is essential.

## Conclusion

To improve upon stagnated recovery rates in eating disorder treatment, it is critical that field move beyond habitual static application of treatment ‘packages’ such as CBT, and towards deconstructing and understanding how these treatments work. The current study represents a significant step towards this goal, being the first to investigate the causal structures responsible for change in a CBT program using causal discovery analysis for a mental health condition. Contrary to the hypothesised mechanisms outlined in the CBT theory for eating disorders, this study provides empirical support for reductions in dietary restraint as the primary driver of subsequent behavioural and cognitive change, including modification in shape and weight concerns. Further research using larger samples, more diagnostically diverse clinical populations, and potentially datasets capturing a broader range of underlying therapeutic drivers will assist in gaining a fuller understanding of the action of CBT.

## Ethics approval and consent to participate

This study has been approved by a Human Research Ethics Review Board (HREC # X18-0486). Participants were required to provide written informed consent prior to being randomised and included in the study.

## Data Availability

All data produced in the present work are contained in the manuscript

## 6. Supplementary Material

**Table S1.**
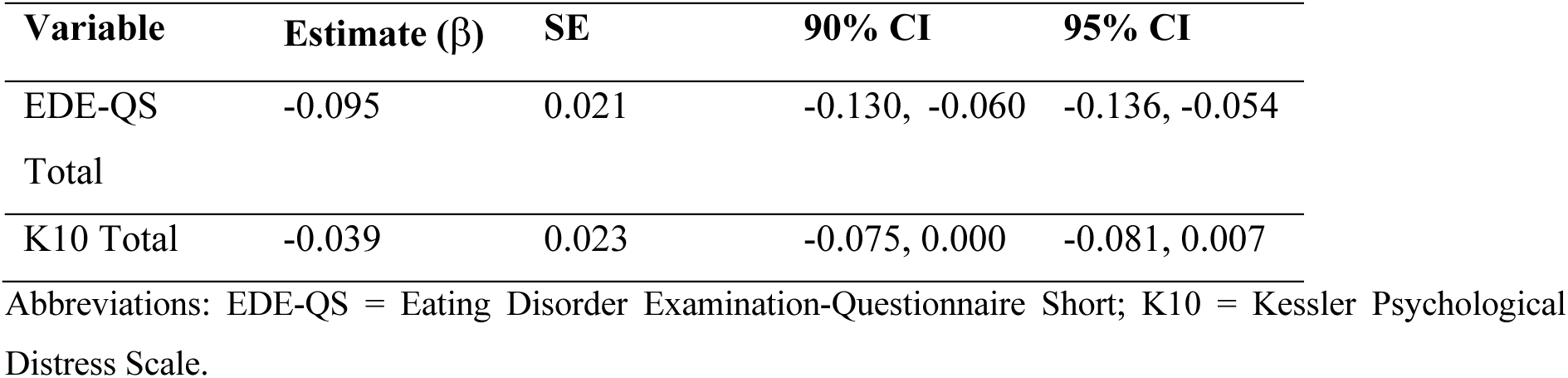
Comparison of Standardised Per-Session Change in EDE-QS and K10 Total Scores Between Trial Arms.

**Table S2.**
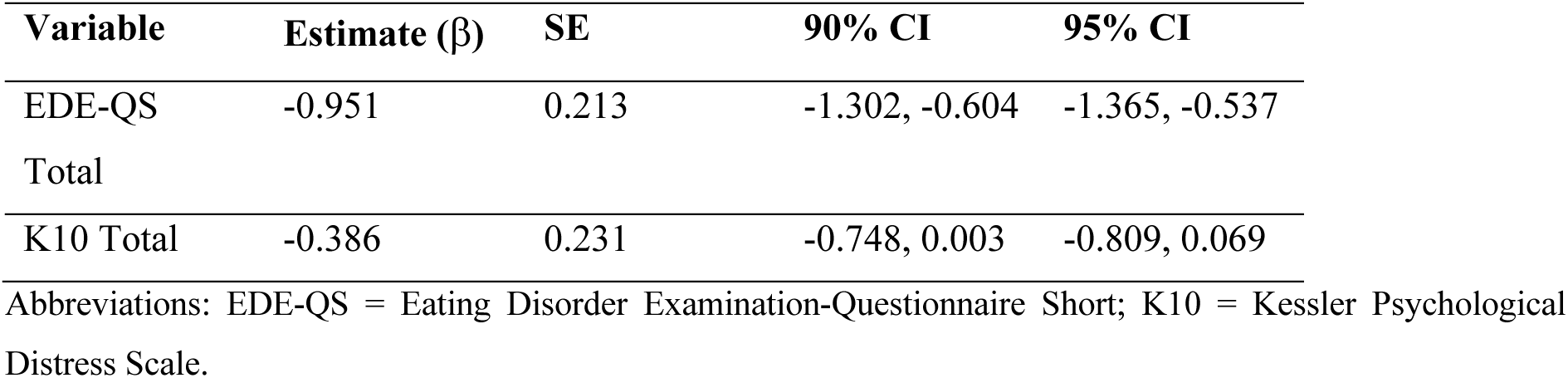
Comparison of Standardised Change at Week 10 in EDE-QS and K10 Total Scores Between Trial Arms.

**Table S3.** Difference between intervention arms and waitlist control for eating disorder and K10 items.

| Item | Estimate | Est.Error | Q2.5 | Q5 | Q95 | Q97.5 |
| --- | --- | --- | --- | --- | --- | --- |
| Eating Restraint | -0.626 | 0.182 | -0.985 | -0.929 | -0.327 | -0.271 |
| Binge | -0.681 | 0.194 | -1.048 | -0.989 | -0.351 | -0.283 |
| Importance Weight | -0.263 | 0.206 | -0.661 | -0.598 | 0.08 | 0.148 |
| Dissatisfaction Weight/Shape | -0.364 | 0.191 | -0.698 | -0.648 | -0.015 | 0.066 |
| Fasting | -0.583 | 0.197 | -0.966 | -0.905 | -0.26 | -0.199 |
| Preoccupation Eating | -0.632 | 0.188 | -1.004 | -0.943 | -0.326 | -0.270 |
| Preoccupation Weight/Shape | -0.457 | 0.207 | -0.860 | -0.798 | -0.121 | -0.061 |
| Fear Weight Gain | -0.604 | 0.226 | -1.027 | -0.965 | -0.225 | -0.158 |
| Desire Weight Loss | -0.431 | 0.251 | -0.914 | -0.842 | -0.019 | 0.050 |
| Vomit/Laxatives | -0.317 | 0.191 | -0.689 | -0.634 | -0.006 | 0.047 |
| Exercise | -0.160 | 0.164 | -0.482 | -0.429 | 0.111 | 0.163 |
| Loss of Control | -0.566 | 0.244 | -1.025 | -0.961 | -0.162 | -0.094 |
| Tired | -0.390 | 0.194 | -0.765 | -0.708 | -0.069 | -0.010 |
| Worthless | -0.335 | 0.156 | -0.638 | -0.585 | -0.075 | -0.019 |
| Nervous | -0.129 | 0.201 | -0.526 | -0.468 | 0.187 | 0.241 |
| Calm | -0.114 | 0.243 | -0.596 | -0.530 | 0.251 | 0.308 |
| Hopeless | -0.144 | 0.205 | -0.572 | -0.505 | 0.172 | 0.227 |
| Restless | -0.236 | 0.173 | -0.578 | -0.523 | 0.047 | 0.100 |
| Sit Still | -0.312 | 0.182 | -0.673 | -0.611 | -0.014 | 0.041 |
| Depressed | -0.228 | 0.244 | -0.684 | -0.627 | 0.145 | 0.202 |
| Effort | -0.354 | 0.209 | -0.735 | -0.681 | 0.004 | 0.069 |
| Cheer | -0.280 | 0.199 | -0.652 | -0.598 | 0.060 | 0.125 |

**Table S4.** Summarised group-by-time comparisons for the pre-post-followup analysis.

| Item | Timepoint | Estimate | Est.Error | Q2.5 | Q5 | Q50 | Q95 | Q97.5 |
| --- | --- | --- | --- | --- | --- | --- | --- | --- |
| Fear Weight Gain | Follow-up | -0.968 | 0.257 | -1.469 | -1.392 | -0.966 | -0.556 | -0.492 |
|  | Post-treatment | -0.618 | 0.254 | -1.099 | -1.041 | -0.587 | -0.241 | -0.185 |
| Desire Weight Loss | Follow-up | -0.832 | 0.204 | -1.250 | -1.183 | -0.824 | -0.514 | -0.459 |
|  | Post-treatment | -0.772 | 0.311 | -1.285 | -1.224 | -0.836 | -0.288 | -0.235 |
| Binge Days | Follow-up | -0.540 | 0.233 | -1.015 | -0.942 | -0.528 | -0.177 | -0.114 |
|  | Post-treatment | -0.434 | 0.206 | -0.886 | -0.825 | -0.406 | -0.136 | -0.091 |
| Vomit | Follow-up | -0.504 | 0.250 | -0.976 | -0.913 | -0.495 | -0.108 | -0.045 |
|  | Post-treatment | -0.513 | 0.158 | -0.818 | -0.770 | -0.514 | -0.251 | -0.200 |
| Laxatives | Follow-up | -0.423 | 0.267 | -1.036 | -0.943 | -0.377 | -0.055 | 0.004 |
|  | Post-treatment | -0.202 | 0.229 | -0.589 | -0.538 | -0.229 | 0.206 | 0.271 |
| Exercise | Follow-up | -0.301 | 0.194 | -0.685 | -0.619 | -0.302 | 0.019 | 0.085 |
|  | Post-treatment | -0.344 | 0.177 | -0.678 | -0.628 | -0.348 | -0.046 | 0.012 |
| Eating Restraint | Follow-up | -0.487 | 0.186 | -0.845 | -0.789 | -0.487 | -0.180 | -0.122 |
|  | Post-treatment | -0.572 | 0.197 | -0.963 | -0.907 | -0.559 | -0.267 | -0.223 |
| Importance Weight | Follow-up | -0.678 | 0.319 | -1.346 | -1.258 | -0.632 | -0.224 | -0.158 |
|  | Post-treatment | -0.453 | 0.197 | -0.814 | -0.759 | -0.464 | -0.108 | -0.042 |
| Dissatisfaction Weight/Shape | Follow-up | -0.612 | 0.223 | -1.037 | -0.974 | -0.615 | -0.247 | -0.188 |
|  | Post-treatment | -0.733 | 0.164 | -1.074 | -1.014 | -0.726 | -0.478 | -0.436 |
| Fasting | Follow-up | -0.594 | 0.252 | -1.070 | -0.993 | -0.607 | -0.155 | -0.077 |
|  | Post-treatment | -0.596 | 0.167 | -0.906 | -0.860 | -0.605 | -0.305 | -0.245 |
| Preoccupation Eating | Follow-up | -0.672 | 0.286 | -1.216 | -1.145 | -0.657 | -0.22 | -0.145 |
|  | Post-treatment | -0.692 | 0.180 | -1.045 | -0.995 | -0.689 | -0.404 | -0.361 |
| Preoccupation Weight/Shape | Follow-up | -0.587 | 0.247 | -1.040 | -0.974 | -0.602 | -0.172 | -0.102 |
|  | Post-treatment | -0.449 | 0.160 | -0.772 | -0.718 | -0.445 | -0.192 | -0.145 |
| Tired | Follow-up | -0.389 | 0.313 | -0.932 | -0.867 | -0.411 | 0.139 | 0.216 |
|  | Post-treatment | -0.445 | 0.235 | -0.884 | -0.823 | -0.450 | -0.053 | 0.008 |
| Worthless | Follow-up | -0.555 | 0.215 | -0.944 | -0.886 | -0.568 | -0.181 | -0.115 |
|  | Post-treatment | -0.319 | 0.164 | -0.633 | -0.583 | -0.324 | -0.041 | 0.014 |
| Nervous | Follow-up | -0.215 | 0.227 | -0.635 | -0.582 | -0.214 | 0.157 | 0.220 |
|  | Post-treatment | -0.330 | 0.292 | -0.851 | -0.798 | -0.268 | 0.086 | 0.139 |
| Calm | Follow-up | -0.129 | 0.21 | -0.517 | -0.459 | -0.14 | 0.233 | 0.301 |
|  | Post-treatment | -0.295 | 0.248 | -0.726 | -0.67 | -0.325 | 0.111 | 0.163 |
| Hopeless | Follow-up | -0.321 | 0.245 | -0.788 | -0.721 | -0.325 | 0.091 | 0.158 |
|  | Post-treatment | -0.435 | 0.241 | -0.836 | -0.785 | -0.467 | 0.002 | 0.067 |
| Restless | Follow-up | -0.239 | 0.260 | -0.715 | -0.649 | -0.244 | 0.185 | 0.256 |
|  | Post-treatment | -0.238 | 0.188 | -0.564 | -0.517 | -0.260 | 0.111 | 0.178 |
| Sit Still | Follow-up | -0.144 | 0.264 | -0.668 | -0.600 | -0.125 | 0.268 | 0.340 |
|  | Post-treatment | -0.483 | 0.194 | -0.845 | -0.792 | -0.492 | -0.155 | -0.096 |
| Depressed | Follow-up | -0.207 | 0.237 | -0.701 | -0.620 | -0.195 | 0.163 | 0.232 |
|  | Post-treatment | -0.428 | 0.193 | -0.782 | -0.733 | -0.437 | -0.106 | -0.051 |
| Effort | Follow-up | -0.291 | 0.387 | -0.926 | -0.856 | -0.34 | 0.412 | 0.505 |
|  | Post-treatment | -0.333 | 0.167 | -0.662 | -0.612 | -0.332 | -0.059 | -0.009 |
| Cheer | Follow-up | -0.204 | 0.339 | -0.742 | -0.681 | -0.249 | 0.426 | 0.511 |
|  | Post-treatment | -0.187 | 0.223 | -0.579 | -0.528 | -0.202 | 0.196 | 0.258 |

**Marginalised edge probabilities for the combined treatment arms (self-help and guided) compared to waitlist control**

**Table S5.** Marginalised edge probabilities for session 2. We highlight cells with edge probabilities greater than 0.5.

| Item | 1 | 2 | 3 | 4 | 5 | 6 | 7 | 8 | 9 | 10 | 11 | 12 | Treatment |
| --- | --- | --- | --- | --- | --- | --- | --- | --- | --- | --- | --- | --- | --- |
| 1 | 0.00 | 0.09 | 0.02 | 0.04 | 0.13 | 0.14 | 0.35 | 0.10 | 0.02 | 0.03 | 0.11 | 0.03 | 0.00 |
| 2 | **0.89** | 0.00 | 0.03 | 0.07 | 0.24 | 0.03 | 0.06 | 0.05 | 0.03 | 0.04 | 0.04 | 0.04 | 0.00 |
| 3 | 0.03 | 0.18 | 0.00 | 0.11 | 0.01 | 0.07 | 0.01 | 0.18 | 0.19 | 0.01 | 0.11 | 0.17 | 0.00 |
| 4 | 0.04 | 0.05 | 0.89 | 0.00 | 0.04 | 0.02 | 0.06 | 0.13 | 0.33 | 0.03 | 0.16 | 0.13 | 0.00 |
| 5 | 0.04 | 0.08 | 0.03 | 0.20 | 0.00 | 0.99 | 0.08 | 0.11 | 0.09 | 0.07 | 0.11 | 0.43 | 0.00 |
| 6 | 0.08 | 0.07 | 0.16 | 0.07 | 0.01 | 0.00 | 0.16 | 0.09 | 0.08 | 0.04 | 0.22 | **0.56** | 0.00 |
| 7 | 0.06 | 0.03 | 0.17 | 0.06 | 0.04 | 0.10 | 0.00 | 0.11 | 0.11 | 0.83 | 0.04 | 0.20 | 0.00 |
| 8 | 0.05 | 0.03 | 0.02 | 0.08 | 0.04 | 0.08 | 0.08 | 0.00 | 0.13 | 0.03 | 0.04 | 0.06 | 0.00 |
| 9 | 0.03 | 0.04 | 0.30 | 0.02 | 0.32 | 0.03 | 0.01 | 0.06 | 0.00 | 0.04 | 0.04 | 0.03 | 0.00 |
| 10 | 0.03 | 0.03 | 0.40 | 0.05 | 0.12 | 0.04 | 0.14 | 0.05 | 0.96 | 0.00 | 0.10 | 0.04 | 0.00 |
| 11 | 0.24 | 0.04 | 0.02 | 0.19 | 0.04 | 0.12 | 0.03 | 0.12 | 0.02 | 0.03 | 0.00 | 0.25 | 0.00 |
| 12 | 0.05 | 0.05 | 0.02 | 0.26 | 0.02 | 0.07 | 0.07 | 0.23 | 0.02 | 0.03 | **0.75** | 0.00 | 0.00 |
| Treatment | 0.09 | 0.06 | 0.04 | 0.18 | 0.06 | 0.05 | 0.10 | 0.07 | 0.20 | 0.05 | 0.06 | 0.06 | 0.00 |

**Table S6.**
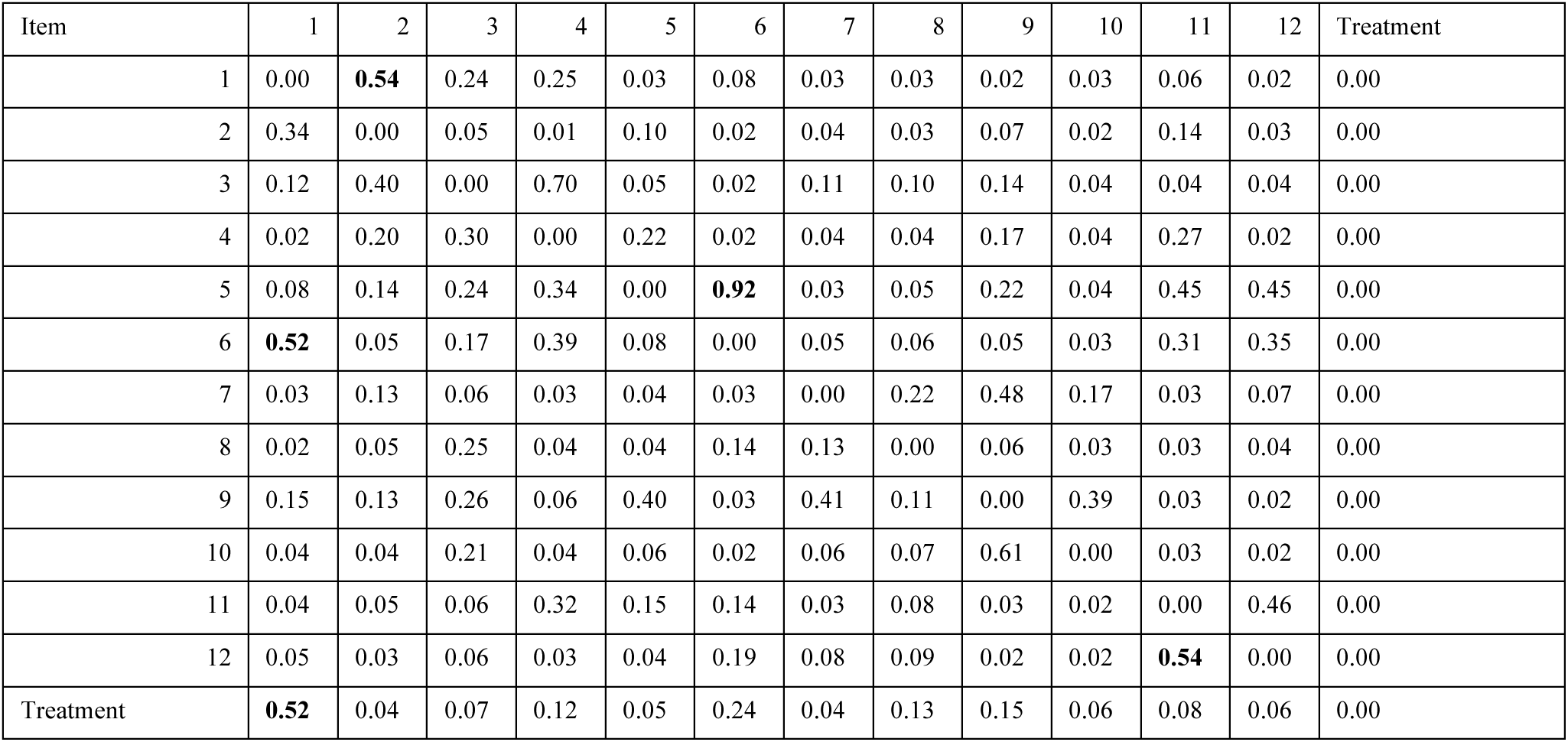
Marginalised edge probabilities for session 3. We highlight cells with edge probabilities greater than 0.5.

**Table S7.** Marginalised edge probabilities for session 4. We highlight cells with edge probabilities greater than 0.5.

| Item | 1 | 2 | 3 | 4 | 5 | 6 | 7 | 8 | 9 | 10 | 11 | 12 | Treatment |
| --- | --- | --- | --- | --- | --- | --- | --- | --- | --- | --- | --- | --- | --- |
| 1 | 0.00 | **0.55** | 0.64 | 0.09 | 0.32 | 0.34 | 0.03 | 0.26 | 0.24 | 0.07 | 0.02 | 0.07 | 0.00 |
| 2 | 0.12 | 0.00 | 0.21 | 0.11 | 0.08 | 0.03 | 0.08 | 0.07 | 0.06 | 0.02 | 0.02 | 0.04 | 0.00 |
| 3 | 0.09 | 0.14 | 0.00 | 0.77 | 0.07 | 0.05 | 0.03 | 0.20 | 0.17 | 0.31 | 0.19 | 0.15 | 0.00 |
| 4 | 0.01 | 0.03 | 0.23 | 0.00 | 0.20 | 0.02 | 0.02 | 0.19 | 0.04 | 0.24 | 0.43 | 0.04 | 0.00 |
| 5 | 0.22 | 0.05 | 0.13 | 0.71 | 0.00 | 0.32 | 0.26 | 0.21 | 0.12 | 0.06 | 0.07 | 0.25 | 0.00 |
| 6 | 0.32 | 0.04 | 0.01 | 0.03 | 0.42 | 0.00 | 0.04 | 0.24 | 0.18 | 0.02 | 0.24 | 0.25 | 0.00 |
| 7 | 0.01 | 0.06 | 0.05 | 0.03 | 0.08 | 0.02 | 0.00 | 0.03 | 0.06 | 0.19 | 0.01 | 0.02 | 0.00 |
| 8 | 0.01 | 0.04 | 0.05 | 0.04 | 0.03 | 0.03 | 0.25 | 0.00 | 0.24 | 0.08 | 0.04 | 0.06 | 0.00 |
| 9 | 0.02 | 0.03 | 0.13 | 0.02 | 0.05 | 0.05 | **0.59** | 0.12 | 0.00 | 0.29 | 0.03 | 0.03 | 0.00 |
| 10 | 0.05 | 0.03 | 0.32 | 0.05 | 0.05 | 0.02 | 0.25 | 0.06 | 0.71 | 0.00 | 0.03 | 0.06 | 0.00 |
| 11 | 0.01 | 0.03 | 0.07 | 0.09 | 0.12 | 0.10 | 0.02 | 0.05 | 0.03 | 0.06 | 0.00 | **0.50** | 0.00 |
| 12 | 0.04 | 0.03 | 0.03 | 0.04 | 0.24 | 0.74 | 0.07 | 0.08 | 0.03 | 0.03 | **0.50** | 0.00 | 0.00 |
| Treatment | **0.82** | **0.55** | 0.11 | 0.09 | 0.08 | 0.04 | 0.14 | 0.10 | 0.08 | 0.08 | 0.04 | 0.07 | 0.00 |

**Table S8.** Marginalised edge probabilities for session 5. We highlight cells with edge probabilities greater than 0.5.

| Node | 1 | 2 | 3 | 4 | 5 | 6 | 7 | 8 | 9 | 10 | 11 | 12 | Treatment |
| --- | --- | --- | --- | --- | --- | --- | --- | --- | --- | --- | --- | --- | --- |
| 1 | 0.00 | **0.63** | 0.12 | 0.20 | 0.05 | 0.31 | 0.04 | **0.64** | 0.03 | 0.06 | 0.05 | 0.07 | 0.00 |
| 2 | 0.10 | 0.00 | 0.05 | 0.02 | 0.05 | 0.02 | 0.03 | 0.03 | 0.05 | 0.03 | 0.03 | 0.03 | 0.00 |
| 3 | 0.06 | 0.06 | 0.00 | 0.21 | 0.04 | 0.02 | 0.07 | 0.03 | 0.23 | 0.12 | 0.02 | 0.04 | 0.00 |
| 4 | 0.15 | 0.04 | **0.79** | 0.00 | 0.49 | 0.16 | 0.07 | 0.17 | **0.61** | 0.04 | 0.36 | 0.08 | 0.00 |
| 5 | 0.14 | 0.12 | 0.07 | 0.18 | 0.00 | 0.54 | 0.16 | 0.18 | 0.08 | 0.08 | 0.20 | 0.06 | 0.00 |
| 6 | **0.59** | 0.30 | 0.06 | 0.03 | 0.43 | 0.00 | 0.18 | 0.14 | 0.03 | 0.03 | 0.38 | 0.43 | 0.00 |
| 7 | 0.02 | 0.13 | 0.05 | 0.03 | 0.10 | 0.03 | 0.00 | 0.14 | 0.49 | 0.08 | 0.02 | 0.06 | 0.00 |
| 8 | 0.05 | 0.05 | 0.09 | 0.08 | 0.02 | 0.11 | 0.29 | 0.00 | 0.04 | 0.09 | 0.02 | 0.11 | 0.00 |
| 9 | 0.01 | 0.09 | 0.30 | 0.21 | 0.11 | 0.04 | 0.46 | 0.05 | 0.00 | 0.72 | 0.02 | 0.05 | 0.00 |
| 10 | 0.11 | 0.06 | 0.07 | 0.03 | 0.04 | 0.03 | 0.09 | 0.03 | 0.28 | 0.00 | 0.10 | 0.05 | 0.00 |
| 11 | 0.03 | 0.07 | 0.11 | 0.64 | 0.16 | 0.07 | 0.03 | 0.08 | 0.04 | 0.08 | 0.00 | **0.70** | 0.00 |
| 12 | 0.03 | 0.11 | 0.13 | 0.02 | 0.14 | 0.49 | 0.03 | 0.08 | 0.04 | 0.05 | 0.30 | 0.00 | 0.00 |
| Treatment | **0.77** | 0.19 | 0.24 | 0.12 | 0.07 | 0.19 | 0.10 | 0.12 | 0.20 | 0.08 | 0.06 | 0.12 | 0.00 |

**Table S9.** Marginalised edge probabilities for session 6. We highlight cells with edge probabilities greater than 0.5.

|  | 1 | 2 | 3 | 4 | 5 | 6 | 7 | 8 | 9 | 10 | 11 | 12 | Treatment |
| --- | --- | --- | --- | --- | --- | --- | --- | --- | --- | --- | --- | --- | --- |
| 1 | 0.00 | 0.36 | 0.67 | 0.13 | 0.11 | 0.16 | 0.04 | 0.23 | 0.30 | 0.10 | 0.06 | 0.05 | 0.00 |
| 2 | 0.03 | 0.00 | 0.03 | 0.02 | 0.17 | 0.02 | 0.23 | 0.20 | 0.21 | 0.09 | 0.06 | 0.03 | 0.00 |
| 3 | 0.01 | 0.08 | 0.00 | 0.53 | 0.06 | 0.04 | 0.15 | 0.06 | 0.03 | 0.09 | 0.12 | 0.27 | 0.00 |
| 4 | 0.01 | 0.02 | 0.47 | 0.00 | 0.36 | 0.02 | 0.12 | 0.07 | 0.12 | 0.02 | 0.37 | 0.13 | 0.00 |
| 5 | **0.57** | 0.06 | 0.05 | 0.49 | 0.00 | 0.71 | 0.05 | 0.14 | 0.04 | 0.07 | 0.07 | 0.29 | 0.00 |
| 6 | 0.28 | 0.03 | 0.37 | 0.20 | 0.21 | 0.00 | 0.05 | 0.06 | 0.13 | 0.04 | 0.52 | 0.83 | 0.00 |
| 7 | 0.05 | 0.09 | 0.06 | 0.03 | 0.05 | 0.02 | 0.00 | **0.52** | 0.04 | 0.07 | 0.13 | 0.04 | 0.00 |
| 8 | 0.06 | 0.43 | 0.02 | 0.03 | 0.02 | 0.02 | 0.07 | 0.00 | 0.02 | 0.06 | 0.03 | 0.05 | 0.00 |
| 9 | 0.14 | 0.38 | 0.48 | 0.24 | 0.13 | 0.25 | 0.49 | 0.16 | 0.00 | 0.46 | 0.06 | 0.05 | 0.00 |
| 10 | 0.30 | 0.05 | 0.24 | 0.05 | 0.10 | 0.05 | 0.45 | 0.15 | 0.54 | 0.00 | 0.03 | 0.07 | 0.00 |
| 11 | 0.01 | 0.02 | 0.03 | 0.18 | 0.01 | 0.06 | 0.05 | 0.04 | 0.05 | 0.02 | 0.00 | 0.37 | 0.00 |
| 12 | 0.01 | 0.02 | 0.03 | 0.03 | 0.04 | 0.15 | 0.05 | 0.07 | 0.01 | 0.02 | 0.45 | 0.00 | 0.00 |
| Treatment | **0.94** | 0.20 | 0.04 | 0.04 | 0.10 | 0.06 | 0.04 | 0.10 | 0.50 | 0.34 | 0.04 | 0.19 | 0.00 |

**Table S10.**
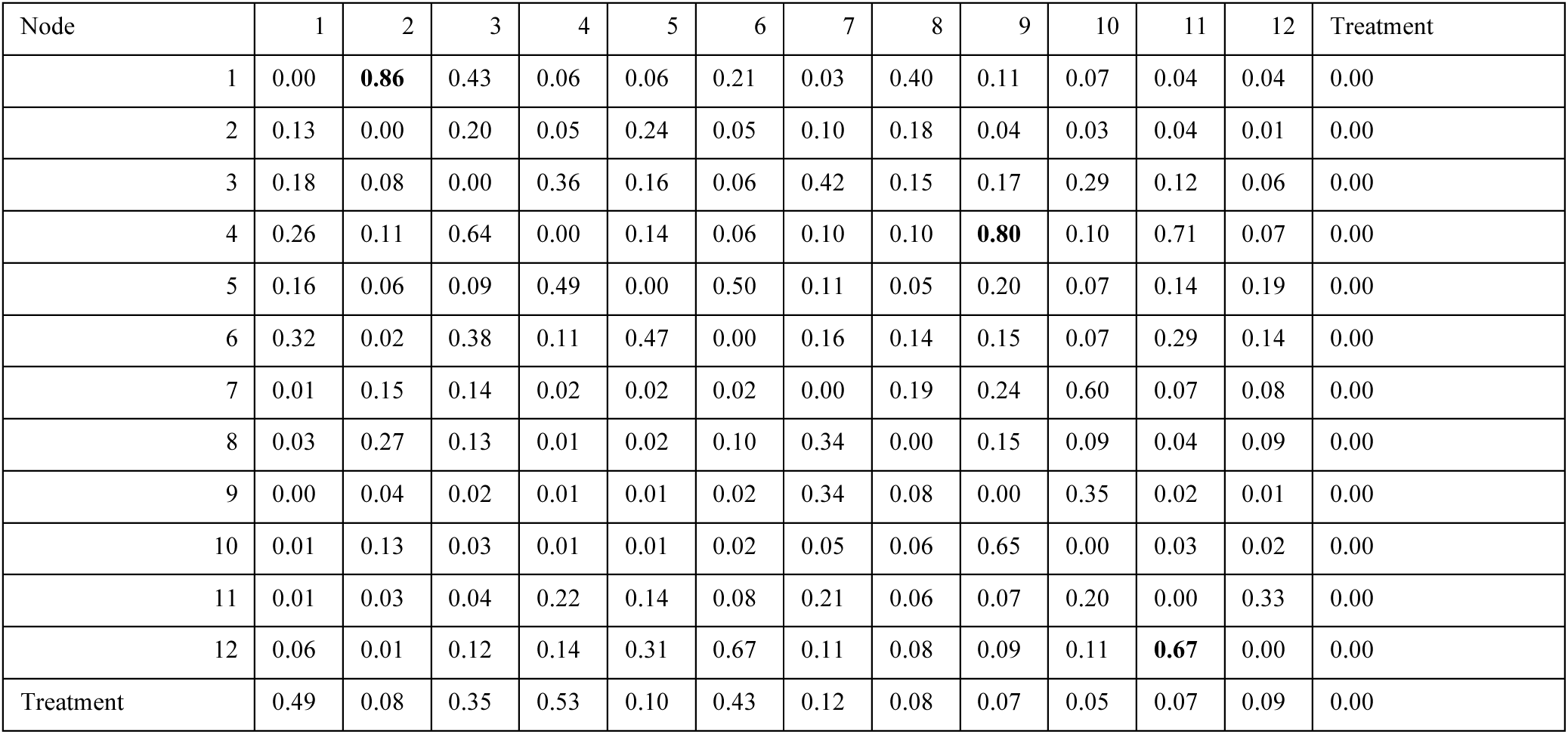
Marginalised edge probabilities for session 7. We highlight cells with edge probabilities greater than 0.5.

**Table S11.** Marginalised edge probabilities for session 8. We highlight cells with edge probabilities greater than 0.5.

| Node | 1 | 2 | 3 | 4 | 5 | 6 | 7 | 8 | 9 | 10 | 11 | 12 | Treatment |
| --- | --- | --- | --- | --- | --- | --- | --- | --- | --- | --- | --- | --- | --- |
| 1 | 0.00 | 0.33 | 0.06 | 0.25 | 0.29 | 0.07 | 0.02 | 0.09 | 0.10 | 0.06 | 0.03 | 0.03 | 0.00 |
| 2 | 0.30 | 0.00 | 0.04 | 0.02 | 0.10 | 0.02 | 0.13 | 0.03 | 0.03 | 0.09 | 0.05 | 0.02 | 0.00 |
| 3 | 0.10 | 0.09 | 0.00 | 0.61 | 0.23 | 0.06 | 0.07 | 0.12 | 0.19 | 0.08 | 0.04 | 0.09 | 0.00 |
| 4 | 0.36 | 0.10 | 0.39 | 0.00 | 0.31 | 0.03 | 0.03 | 0.05 | 0.25 | 0.07 | 0.26 | 0.16 | 0.00 |
| 5 | 0.16 | 0.21 | 0.19 | 0.37 | 0.00 | 0.37 | 0.05 | 0.09 | 0.04 | 0.11 | 0.08 | 0.13 | 0.00 |
| 6 | 0.26 | 0.05 | 0.10 | 0.17 | 0.63 | 0.00 | 0.08 | 0.07 | 0.02 | 0.07 | 0.29 | 0.28 | 0.00 |
| 7 | 0.06 | 0.28 | 0.04 | 0.03 | 0.03 | 0.03 | 0.00 | 0.27 | 0.11 | 0.49 | 0.10 | 0.13 | 0.00 |
| 8 | 0.03 | 0.05 | 0.16 | 0.02 | 0.04 | 0.08 | 0.56 | 0.00 | 0.07 | 0.09 | 0.12 | 0.20 | 0.00 |
| 9 | 0.04 | 0.06 | 0.37 | 0.09 | 0.03 | 0.19 | 0.48 | 0.18 | 0.00 | **0.65** | 0.25 | 0.07 | 0.00 |
| 10 | 0.02 | 0.12 | 0.31 | 0.02 | 0.14 | 0.09 | 0.15 | 0.09 | 0.35 | 0.00 | 0.19 | 0.04 | 0.00 |
| 11 | 0.03 | 0.04 | 0.04 | 0.29 | 0.06 | 0.28 | 0.14 | 0.08 | 0.13 | 0.19 | 0.00 | 0.46 | 0.00 |
| 12 | 0.16 | 0.03 | 0.09 | 0.05 | 0.05 | 0.41 | 0.16 | 0.11 | 0.05 | 0.07 | **0.54** | 0.00 | 0.00 |
| Treatment | 0.32 | 0.21 | 0.29 | 0.09 | 0.08 | 0.09 | 0.05 | 0.11 | 0.65 | 0.14 | 0.12 | 0.21 | 0.00 |

**Table S12.** Marginalised edge probabilities for session 9. We highlight cells with edge probabilities greater than 0.5.

| Node | 1 | 2 | 3 | 4 | 5 | 6 | 7 | 8 | 9 | 10 | 11 | 12 | Treatment |
| --- | --- | --- | --- | --- | --- | --- | --- | --- | --- | --- | --- | --- | --- |
| 1 | 0.00 | 0.43 | 0.24 | 0.04 | 0.20 | 0.21 | 0.02 | 0.14 | 0.08 | 0.04 | 0.04 | 0.01 | 0.00 |
| 2 | 0.42 | 0.00 | 0.03 | 0.01 | 0.06 | 0.19 | 0.15 | 0.04 | 0.05 | 0.02 | 0.07 | 0.02 | 0.00 |
| 3 | 0.44 | 0.44 | 0.00 | 0.26 | 0.05 | 0.06 | 0.13 | 0.03 | 0.32 | 0.15 | 0.05 | 0.04 | 0.00 |
| 4 | 0.23 | 0.06 | 0.74 | 0.00 | 0.10 | 0.02 | 0.03 | 0.04 | 0.48 | 0.05 | 0.22 | 0.05 | 0.00 |
| 5 | 0.17 | 0.03 | 0.04 | 0.23 | 0.00 | 0.24 | 0.03 | 0.05 | 0.06 | 0.03 | 0.05 | 0.10 | 0.00 |
| 6 | 0.28 | 0.06 | 0.08 | 0.14 | **0.76** | 0.00 | 0.11 | 0.07 | 0.04 | 0.05 | 0.21 | 0.27 | 0.00 |
| 7 | 0.02 | 0.18 | 0.50 | 0.26 | 0.06 | 0.09 | 0.00 | 0.15 | 0.33 | 0.42 | 0.05 | 0.12 | 0.00 |
| 8 | 0.08 | 0.11 | 0.04 | 0.05 | 0.08 | 0.09 | 0.34 | 0.00 | 0.11 | 0.23 | 0.11 | 0.07 | 0.00 |
| 9 | 0.01 | 0.06 | 0.12 | 0.07 | 0.04 | 0.02 | 0.10 | 0.10 | 0.00 | 0.48 | 0.35 | 0.04 | 0.00 |
| 10 | 0.02 | 0.10 | 0.09 | 0.03 | 0.03 | 0.02 | 0.27 | 0.08 | 0.52 | 0.00 | 0.20 | 0.01 | 0.00 |
| 11 | 0.02 | 0.03 | 0.04 | 0.25 | 0.07 | 0.06 | 0.03 | 0.11 | 0.06 | 0.20 | 0.00 | 0.09 | 0.00 |
| 12 | 0.05 | 0.03 | 0.25 | 0.70 | 0.20 | 0.72 | 0.21 | 0.06 | 0.06 | 0.06 | 0.91 | 0.00 | 0.00 |
| Treatment | 0.36 | 0.31 | 0.07 | 0.05 | 0.11 | 0.40 | 0.04 | 0.25 | 0.19 | 0.10 | 0.06 | 0.28 | 0.00 |

**Table S13.** Marginalised edge probabilities for session 10. We highlight cells with edge probabilities greater than 0.5.

| Node | 1 | 2 | 3 | 4 | 5 | 6 | 7 | 8 | 9 | 10 | 11 | 12 | Treatment |
| --- | --- | --- | --- | --- | --- | --- | --- | --- | --- | --- | --- | --- | --- |
| 1 | 0.00 | 0.26 | 0.37 | 0.25 | 0.09 | 0.39 | 0.24 | 0.22 | 0.07 | 0.04 | 0.06 | 0.03 | 0.00 |
| 2 | 0.23 | 0.00 | 0.15 | 0.03 | 0.05 | 0.08 | 0.05 | 0.10 | 0.18 | 0.03 | 0.34 | 0.02 | 0.00 |
| 3 | 0.19 | 0.25 | 0.00 | 0.44 | 0.08 | 0.08 | 0.02 | 0.14 | 0.16 | 0.10 | 0.06 | 0.06 | 0.00 |
| 4 | 0.09 | 0.04 | **0.56** | 0.00 | 0.27 | 0.03 | 0.01 | 0.07 | 0.16 | 0.09 | 0.07 | 0.15 | 0.00 |
| 5 | 0.09 | 0.03 | 0.14 | 0.13 | 0.00 | 0.42 | 0.02 | 0.11 | 0.10 | 0.03 | 0.13 | 0.18 | 0.00 |
| 6 | **0.53** | 0.07 | 0.31 | 0.11 | 0.57 | 0.00 | 0.02 | 0.07 | 0.16 | 0.05 | 0.15 | 0.42 | 0.00 |
| 7 | 0.14 | 0.33 | 0.04 | 0.03 | 0.23 | 0.12 | 0.00 | 0.29 | 0.48 | 0.07 | 0.11 | 0.08 | 0.00 |
| 8 | 0.05 | 0.04 | 0.08 | 0.03 | 0.04 | 0.03 | 0.25 | 0.00 | 0.03 | 0.21 | 0.04 | 0.18 | 0.00 |
| 9 | 0.03 | 0.17 | 0.26 | 0.09 | 0.13 | 0.09 | 0.51 | 0.13 | 0.00 | 0.76 | 0.09 | 0.08 | 0.00 |
| 10 | 0.02 | 0.03 | 0.12 | 0.12 | 0.03 | 0.04 | 0.08 | 0.19 | 0.24 | 0.00 | 0.04 | 0.05 | 0.00 |
| 11 | 0.12 | 0.25 | 0.16 | 0.21 | 0.36 | 0.51 | 0.04 | 0.09 | 0.20 | 0.07 | 0.00 | **0.64** | 0.00 |
| 12 | 0.05 | 0.02 | 0.26 | 0.76 | 0.40 | 0.49 | 0.03 | 0.08 | 0.12 | 0.05 | 0.26 | 0.00 | 0.00 |
| Treatment | **0.59** | 0.40 | 0.10 | 0.06 | 0.16 | 0.08 | 0.07 | 0.08 | 0.39 | 0.17 | 0.12 | 0.12 | 0.00 |

